# Toward Equity in Health: A Scoping Review Protocol for Fifteen Years of Progress and Gaps in Disability-Inclusive Healthcare (2010–2025)

**DOI:** 10.1101/2025.08.31.25334801

**Authors:** Lufuno Makhado, Thendo Gertie Makhado

## Abstract

**Background:** Persons with disabilities (PWDs) experience persistent disparities in accessing equitable and effective healthcare, despite the existence of global human rights frameworks. Structural, financial, attitudinal, and systemic barriers continue to impede their full inclusion in healthcare systems. Over the past fifteen years, frameworks such as the UN Convention on the Rights of Persons with Disabilities (CRPD) have sought to promote inclusive health reforms. However, implementation remains uneven, and a comprehensive synthesis of interventions is lacking.

**Objective:** This scoping review aims to map and synthesise global literature (2010– 2025) on disability-inclusive healthcare systems. It focuses on structural competency, universal design, telehealth, and community-based rehabilitation (CBR), identifying key barriers and facilitators.

**Methods:** The review will follow Arksey and O’Malley’s scoping review methodology, incorporating Joanna Briggs Institute (JBI) and PRISMA-ScR guidance. Literature will be identified through searches in PubMed, Scopus, Web of Science, CINAHL, and Google Scholar. Eligibility will be guided by the Population–Concept– Context (PCC) framework, with studies screened and charted independently by two reviewers. Thematic synthesis and evidence mapping will be used to analyse findings. No meta-analysis will be conducted due to expected heterogeneity.

**Expected Outcomes:** The review is anticipated to categorise major systemic barriers (e.g., physical inaccessibility, provider bias, financial exclusion) and identify inclusive interventions aligned with CRPD principles. These insights will support health policy reform and training in disability-responsive care. This review addresses a key gap by systematically synthesising global evidence on disability-inclusive healthcare interventions with a specific focus on structural competency, a lens rarely applied in previous reviews.

**Ethics and Registration:** No ethical approval is required. This protocol is registered with the Open Science Framework (OSF) https://doi.org/10.17605/OSF.IO/PWC92.

## Introduction

Healthcare accessibility is a critical component of achieving global health equity. The necessity of this accessibility is underscored by the fact that, despite the existence of established human rights frameworks [1,2], people with disabilities (PWDs) continue to experience significantly higher rates of morbidity and premature mortality, along with poorer health outcomes [3,4]. With over 1.3 billion individuals living with disabilities worldwide [5,6], it is imperative to address the systemic barriers they face in accessing healthcare services.

The academic discourse surrounding health disparities has evolved over time. Initially focused on impairment models, scholarship is increasingly examining structural determinants of inequity [7]. This shift emphasises that inequities are not simply the result of individual physical limitations but are deeply rooted in societal structures that create and perpetuate disadvantage. In this context, the concept of structural competency emerges as a transformative framework aimed at addressing these systemic obstacles [8,9]. By equipping healthcare professionals with the skills to recognise and challenge these structural factors, we can work towards creating a more equitable healthcare system.

The urgency of addressing these issues has been further amplified by the COVID-19 pandemic, which has exposed and exacerbated existing inequities within healthcare systems [10,11]. Vulnerable populations, particularly PWDs, have faced increased challenges in accessing timely and adequate care during this crisis. This highlights the critical need for targeted interventions and thorough evaluations of existing healthcare frameworks. Despite growing attention, no scoping review to date has synthesised evidence on disability-inclusive healthcare globally using the structural competency framework. This review will therefore provide critical insights to guide policy, practice, and research.

## Materials and Methods

### Aim

Acknowledging the significance of understanding and addressing barriers faced by PWDs, this review synthesises literature from 2010 to 2025. It aims to identify barriers to healthcare accessibility, evaluate the effectiveness of interventions to overcome these challenges, and assess the application of structural competency in different healthcare settings. This study seeks to inform policy and practice, contributing to more equitable healthcare outcomes for individuals with disabilities. Additionally, it aims to map the extent and nature of the literature on disability-inclusive healthcare systems.

### Research Question

The Population–Concept–Context (PCC) framework was used to develop a focused and methodologically robust research question that aligns with the scoping review’s objective: to identify, map, and synthesise systemic barriers and promising interventions for disability-inclusive healthcare globally.

Review Question:

*What are the key barriers to and facilitators of disability-inclusive healthcare systems globally, and which interventions have been implemented or proposed between 2010 and 2025 to improve equitable access for persons with disabilities?*

This question is deliberately broad in scope, allowing for exploration of both structural-level reforms and community-based solutions. The PCC framework ensured comprehensiveness without loss of conceptual clarity, particularly in distinguishing systemic interventions from individual clinical measures.

### Study Registration subsection

This scoping review protocol has been prospectively registered with the Open Science Framework (OSF) at https://doi.org/10.17605/OSF.IO/PWC92. The registration includes the full protocol, eligibility criteria, and planned methods to ensure transparency and reproducibility.

### Design and Setting

The study design is guided by Arksey and O’Malley’s five-stage framework and updated Joanna Briggs Institute (JBI) guidance [12]. The setting includes global literature in English from 2010 to 2025.

#### Eligibility Criteria: PCC Framework

This scoping review utilised the PCC framework recommended by the JBI to guide the development of inclusion criteria and evidence synthesis strategy [12].

The Population focuses on PWDs across the lifespan and across all types of impairments. The Concept captures disability-inclusive healthcare systems and the interventions designed to improve access and equity. The Context encompasses global healthcare systems, public, private, and community-based, spanning both high- and low-resource settings, within the time frame of 2010 to 2025.

This structured approach will allow for a broad yet targeted inquiry into systemic healthcare barriers and inclusive interventions, ensuring the review captures both policy and practical innovations across settings.

| Component | Criteria |
| --- | --- |
| **Population (P)** | Persons with disabilities (PWDs), including children, adults, and older adults, regardless of disability type (physical, sensory, cognitive, psychosocial, etc.) |
| **Concept (C)** | Disability-inclusive healthcare systems and interventions, including but not limited to: universal design, structural competency training, telehealth and assistive technologies, CRPD-aligned policy reforms, and community-based rehabilitation (CBR) |
| **Context (C)** | Global healthcare settings, including public and private health systems, across all income settings. The review spans literature published between 2010 and 2025, in English only |

### Search Strategy

The following advanced search strategy will be employed to comprehensively explore the topic:

#### Databases searched include

- PubMed
- Scopus
- Web of Science
- CINAHL
- Google Scholar (top 200 results)
- Organisational websites (including but not limited to, WHO, UN, World Bank)
- Disability advocacy group repositories

A structured search strategy was developed using Boolean operators and a combination of relevant keywords and phrases to maximise the breadth and depth of the search. Both peer-reviewed and grey literature will be included in this review to capture policy reports, NGO publications, government documents, and other relevant non-academic sources. Grey literature will be searched through databases such as Google Scholar, organisational websites, and disability advocacy group repositories to ensure inclusion of community-driven innovations. The terms were organised into three main categories:

#### Disability Context, which will include

- disability
- “persons with disabilities”
- PWD

#### Healthcare Accessibility Themes

- ***“***inclusive healthcare”
- “healthcare accessibility”
- “universal design”
- “health equity”

#### “Education and Systemic Framework

- “structural competency”
- “medical education”
- “health systems”

The following search strings will be used in this review, combining the three categories through Boolean logic:

(disability OR “persons with disabilities” OR PWD)

AND

(“inclusive healthcare” OR “healthcare accessibility” OR “universal design” OR “health equity”)

AND

(“structural competency” OR “medical education” OR “health systems”)

The following filters will be applied to ensure that the scoping review search strategy aligns with the study’s eligibility criteria. Specific filters have been implemented to refine the search results that will be applicable.

- publication date range
- type of publication (e.g., peer-reviewed articles, systematic reviews)
- language (e.g., English)

The process to be followed includes that the search results will be systematically reviewed, starting with the titles and abstracts that will be screened for relevance, and full texts will be evaluated for inclusion based on predetermined criteria. All relevant studies will then be extracted and analysed for themes related to inclusive healthcare practices and education concerning people with disabilities (See *Data Charting and management section*). This rigorous strategy ensured a thorough investigation of the literature, capturing diverse perspectives on the intersection of disability and healthcare accessibility.

### Study Selection

The selection of studies will follow the PRISMA-ScR guidelines to ensure transparency and reproducibility [13,14]. All citations retrieved from the database searches (PubMed, Scopus, Web of Science, CINAHL, and Google Scholar) will be imported into a reference management tool (in this case, EndNote) for de-duplication.

Following this, titles and abstracts will be exported to a spreadsheet for initial screening. Two reviewers (LM and TGM) will independently screen the titles and abstracts against the predefined inclusion and exclusion criteria guided by the PCC framework through the use of Covidence Software. Any article deemed potentially relevant or where eligibility is unclear will be retrieved in full for further assessment.

The full-text screening will also be conducted independently by both reviewers. Eligibility will be determined based on the alignment with the review’s population (PWDs), concept (inclusive healthcare interventions), and context (healthcare systems globally, 2010–2025). Any disagreements at this stage will be resolved through discussion and consensus; if consensus cannot be reached, a third reviewer will be consulted. Reasons for excluding studies at the full-text screening stage will be recorded in a log and summarised in the final review report. The study selection process will be illustrated using a PRISMA-ScR flow diagram in the completed review [13,14]. Additionally, reference lists of included studies will be hand-searched to identify further eligible articles. The entire process will be visualised using a PRISMA-ScR flow diagram [13,14].

### Data Charting and Management

Data will be extracted from all included studies using a standardised data charting form developed and piloted by the review team. The following data items will be captured:

- Author(s) and year of publication
- Study location/region
- Study design and methodology
- Population characteristics (e.g., age, disability type)
- Type of healthcare setting
- Identified barriers (e.g., physical, attitudinal, systemic, financial)
- Type and description of interventions (e.g., universal design, telehealth, policy reform, CBR)
- Outcomes or impacts reported
- Alignment with CRPD principles or structural competency frameworks

Two reviewers will independently chart data from included studies. The data charting form may be iteratively updated as familiarity with the literature increases. Any discrepancies will be resolved by discussion or the involvement of a third reviewer. In addition to data charting, the quality of included studies will be assessed using the Mixed Methods Appraisal Tool (MMAT) [15], which allows appraisal across qualitative, quantitative, and mixed-methods designs. This step is essential given the heterogeneity anticipated in study designs. Quality assessment will not be used to exclude studies but will inform the interpretation of findings and highlight evidence strength and gaps.

### Expected Outcomes and Analysis Plan

The primary outcome of this scoping review will be to identify and categorise the key barriers to disability-inclusive healthcare. These barriers will include physical, financial, attitudinal, and systemic obstacles. Secondary outcomes will involve mapping a variety of interventions that have been designed or implemented to address these barriers. Such interventions may include universal design principles, telehealth and assistive technologies, structural competency training in health professions education, CRPD-aligned policy reforms, and community-based rehabilitation (CBR) approaches.

A thematic synthesis approach will be employed to analyse the extracted data. The data will be organised into key thematic domains based on patterns that emerge across studies. These domains are expected to align with the following categories:

- Nature and type of barrier (e.g., infrastructure, affordability, discrimination)
- Type and scale of intervention (e.g., national policy, community-level programs)
- Outcomes and impacts on healthcare access, quality, and equity for persons with disabilities (PWDs)

Descriptive statistics, such as frequencies and distributions, will be used, where applicable, to summarise the characteristics of the studies and the types of interventions. An evidence map will also be created to visually present the distribution of studies by theme, region, intervention, and population group.

Due to the anticipated heterogeneity of study designs, contexts, and outcome measures, no meta-analysis will be conducted. Instead, the findings will be synthesised narratively and presented in summary tables, figures, and charts to enhance interpretability and utility for policymakers and practitioners. Findings will be synthesised narratively and thematically, with results contextualised according to the methodological quality of included studies as assessed by the MMAT [15].

### Ethical Considerations

As a scoping review, this study involves no human subjects and does not require ethical approval.

### Study Status and Timeline

This scoping review will commence in October 2025, following the publication of the protocol and registration in OSF. All methods and procedures described in this protocol will be followed systematically. Any amendments made during the review process will be documented and updated in the OSF Registries. The current status of this study is at the protocol level. We estimate the following timeline for completion of each phase: A) The search strategy and selection of eligible studies for inclusion in the review will be completed by December 2025; B) Data extraction, charting, and critical appraisal will be completed by February 2026; and C) Analysis, results, and synthesis are expected to be completed by April 2026.

## Discussion

This scoping review protocol outlines a comprehensive and methodologically sound plan for synthesising evidence on disability-inclusive healthcare systems from 2010 to 2025. Utilising the PCC framework and following the JBI/PRISMA-ScR guidelines, this review aims to map the nature, range, and scope of both barriers and interventions that affect healthcare access for persons with disabilities (PWDs) worldwide. The findings of this review are expected to inform health policy, strengthen inclusive healthcare practices, and guide medical education curricula. By applying a structural competency lens, the review has the potential to influence systemic reforms that extend beyond individual-level interventions.

### Strengths and Limitations

This review’s key strength is its alignment with global health equity frameworks, like the CRPD, enhancing its relevance to international policy. It employs rigorous methods, including the Arksey and O’Malley framework, JBI guidance, and OSF registration, ensuring transparency and reproducibility. The use of the PCC framework allows for systematic eligibility determination, while a multi-database search enhances comprehensiveness. Reliability is strengthened through dual independent screening and standardised data charting. The thematic synthesis, combined with evidence mapping, provides depth and breadth, and the focus on structural competency expands disability research beyond individual impairment models. Plans for dissemination, such as open-access publication and plain-language summaries, ensure accessibility and policy relevance.

However, limitations include the restriction to English-language studies, which may exclude important non-English evidence, and a fixed time frame of 2010 to 2025, which risks missing emerging evidence. Additionally, database coverage bias may limit representation from certain regions.

### Dissemination Plans

The findings from the completed scoping review will be effectively communicated through a variety of channels to ensure accessibility for diverse audiences. To begin, a manuscript detailing the review will be submitted to a peer-reviewed, open-access journal that specialises in public health or disability studies. In addition, a plain-language summary will be developed to facilitate understanding among community stakeholders, disability advocacy groups, and policymakers. To further disseminate the information, presentations will be delivered at relevant academic and professional conferences, including the International Conference on Disability Studies scheduled for 2026. Lastly, whenever possible, the findings will also be made available on institutional websites and open-access repositories to maximise their reach and impact.

### Amendments and Protocol Deviations

Any changes to the protocol, including modifications to eligibility criteria, data extraction categories, or synthesis methods, will be thoroughly documented. These updates will be included in the final publication with appropriate justifications. The protocol is registered with OSF, and any significant changes will be promptly reflected in the OSF record to maintain transparency. In cases of project termination or major delays, we will provide clear documentation that explains the rationale behind these decisions, along with any challenges or limitations faced. This approach ensures accountability and clarity in our reporting, even if the review is not completed as initially intended.

## Conclusion

This scoping review protocol outlines a clear approach to mapping global evidence on disability-inclusive healthcare systems from 2010 to 2025. By incorporating both peer-reviewed and grey literature and using the Mixed Methods Appraisal Tool, the review will identify barriers and effective interventions, such as universal design and telehealth. Utilising the structural competency framework, it aims to address systemic inequities. The findings will inform policymakers, healthcare providers, and educators, advancing disability-inclusive health policy and strengthening commitments under the CRPD.

## Data Availability

No datasets were generated or analysed during the current study. All relevant data from this study will be made available upon study completion.

## Authors’ Contributions

**Conceptualisation:** Lufuno Makhado, Thendo Gertie Makhado.

**Investigation:** Lufuno Makhado, Thendo Gertie Makhado.

**Methodology:** Lufuno Makhado, Thendo Gertie Makhado.

**Writing – original draft:** Lufuno Makhado

**Writing – review & editing:** Lufuno Makhado & Thendo Gertie Makhado

## Acknowledgements

We thank the University of Venda for institutional support.

## Supporting Information

- PRISMA-ScR Checklist
- OSF Registration: The full protocol is publicly available at https: https://doi.org/10.17605/OSF.IO/PWC92

